# Decreased serum levels of the inflammaging marker miR-146a are associated with clinical response to tocilizumab in COVID-19 patients

**DOI:** 10.1101/2020.07.11.20151365

**Authors:** Jacopo Sabbatinelli, Angelica Giuliani, Giulia Matacchione, Silvia Latini, Noemi Laprovitera, Giovanni Pomponio, Alessia Ferrarini, Silvia Svegliati Baroni, Marianna Pavani, Marco Moretti, Armando Gabrielli, Antonio Domenico Procopio, Manuela Ferracin, Massimiliano Bonafè, Fabiola Olivieri

## Abstract

Current COVID-19 pandemic poses an unprecedented threat to global health and healthcare systems. The most amount of the death toll is accounted by old people affected by age-related diseases that develop a hyper-inflammatory syndrome. In this regard, we hypothesized that COVID-19 severity may be linked to inflammaging. Here, we examined 30 serum samples from patients enrolled in the clinical trial NCT04315480 assessing the clinical response to a single-dose intravenous infusion of the anti-IL-6 receptor drug Tocilizumab (TCZ) in COVID-19 patients with multifocal interstitial pneumonia.

In these serum samples, as well as in 29 age- and gender-matched healthy control subjects, we assessed a set of microRNAs that regulate inflammaging, i.e. miR-146a-5p, miR-21-5p, and miR-126-3p, which were quantified by RT-PCR and Droplet Digital PCR.

We showed that COVID-19 patients who did not respond to TCZ have lower serum levels of miR-146a-5p after the treatment (p=0.007). Among non-responders, those with the lowest serum levels of miR-146a-5p experienced the most adverse outcome (p=0.008). Our data show that a blood-based biomarker, such as miR-146a-5p, can provide clues about the molecular link between inflammaging and COVID-19 clinical course, thus allowing to better understand the use of biologic drug armory against this worldwide health threat.

## 1. INTRODUCTION

The COVID-19 pandemic caused by the SARS-CoV-2 coronavirus is characterized by a striking age-dependent morbidity and mortality, irrespective of ethnicity (Remuzzi and Remuzzi, 2020; Ruan et al., 2020). In Italy, about 1.1% of COVID-19 patient deaths are attributable to people younger than 50 years, while the death toll of COVID-19 is mainly accounted by people with a median age of 82 years (IQR 74-88). Notably, most of such patients (59.7%) are affected by at least three age-related diseases (Istituto Superiore di Sanità, 2020). Nowadays, whilst the pandemic is still spreading, COVID-19 is emerging as a transmissible age-related lethal disease, at least in western countries.

A substantial proportion of hospitalized COVID-19 patients show a systemic dysregulation of pro-inflammatory cytokines, a condition known as cytokine storm (Zhou et al., 2020). Such hyper-inflammatory response in COVID-19 patients is associated with extensive lung and endothelial cell damage, microvascular dysfunction, and microangiopathy that eventually lead to lung and multiorgan failure (Varga et al., 2020). It is therefore unsurprising that a number of clinical trials have been started to evaluate the effectiveness of anti-cytokine/cytokine receptor antibodies as a treatment for hospitalized COVID-19 patients. Tocilizumab (TCZ), the monoclonal antibody against the interleukin-6 (IL-6) receptor is one of these host-directed therapies (Crisafulli et al., 2020; Liu et al., 2020a; Luo et al., 2020; Xu et al., 2020). Nevertheless, substantial variability in the clinical response of COVID-19 patients to TCZ treatment has been reported, probably due to the contribution of other factors, including age-related biological mechanisms, gender, genetic makeup, disease severity, timing of treatment, and immune activation (Khiali et al., 2020; Wilk et al., 2020). With regard to the latter issue, we have recently proposed that an age-dependent pro-inflammatory status, currently referred to as inflammaging, may facilitate the onset of uncontrolled COVID-19 related hyper-inflammation, particularly in aged people (Bonafè et al., 2020; Storci et al., 2020). Blood-based biomarkers, conceivably linked to the systemic inflammatory state and/or inflammaging, and able to predict the response to TCZ treatment are urgently needed in order to redirect this precious armory to those patients who are likely to benefit at its most. Under this perspective, the assessment of serum/plasma microRNAs (miRNAs) has been emerging as a reliable tool to assess pharmacological interventions in a variety of human diseases, including age-related and infectious diseases (Kong et al., 2012).

In this regard, a set of miRNAs capable of regulating inflammation (also known as inflamma-miRs) such as miR-146a, miR-21-5p, and miR-126-3p, has reported to show age-dependent changes in their blood levels and to be markers of a pro-inflammatory state (i.e. inflammaging) in healthy and non-healthy individuals (Mensà et al., 2019; Olivieri et al., 2013b; Olivieri et al., 2012). These three miRNAs are all able to target molecules belonging to the nuclear factor κB (NF-κB) pathway, a central mediator of inflammation (Mussbacher et al., 2019). Hyper-inflammatory response is associated with poor outcome in COVID-19 patients and shows multiple connections with the thrombotic phenomena that occur in patients affected by severe COVID-19 (Sriram and Insel, 2020). MiR-146a-5p is of particular interest, given its ability to induce a negative feedback loop that restrains the activation of the NF-κB pathway by targeting TNF receptor associated factor 6 (TRAF6) and Interleukin-1 receptor-associated kinase 1 (IRAK1). The pivotal role of miR-146a-5p has been reported in a number of cell types that orchestrate the inflammatory response, including endothelial cells (Mensà et al., 2019; Olivieri et al., 2013a), microglial cells (Liu et al., 2020b), monocytes and macrophages, (Li et al., 2015; Zhou et al., 2019), and adipocytes (Roos et al., 2016). Recently, a reduced expression of miR-146a-5p was observed in a subgroup of patients affected by acute myeloid leukemia with inflammaging and poor outcome (Grants et al., 2020). Therefore, on the basis of the current literature, a decline in miR-146a-5p levels would be expected to unleash the release of inflammatory cytokines, e.g. IL-6.

Other inflammatory miRNAs, including miR-21-5p, take part to the scenario. This microRNA was previously identified as a cell-free and/or exosome-loaded circulating marker of inflammaging and endothelial senescence (Mensà et al., 2020; Olivieri et al., 2012). Even miR-126-3p, a key regulator of endothelial inflammation and angiogenic processes, has been regarded as a mediator of systemic inflammation (reviewed in Zhong et al., 2018).

The aim of this report is to evaluate the levels of circulating inflamma-miR-146a, -21-5p, and -126-3p in COVID-19 patients treated with TCZ and age-matched healthy control subjects, in order to assess whether such biomarkers of inflammaging may be of help in such a dramatic clinical setting.

## 2. MATERIALS AND METHODS

Thirty serum samples from COVID-19 patients enrolled in a Phase 2, open-label, single-arm clinical trial (Clinicaltrials.gov, NCT04315480) were available for miRNA assay. The clinical trial was conducted at Università Politecnica delle Marche, Italy and enrolled 46 patients presenting with virologically confirmed COVID-19, characterized by multifocal interstitial pneumonia confirmed by CT-scan and requiring oxygen therapy. Eligible patients presented with worsening respiratory function, defined as either decline of oxygen saturation (SO2) > 3% or a > 50% decline in PaO2/FiO2 (P/F) ratio in the previous 24h; need to increase FiO2 in order to maintain a stable SO2 or new-onset need of mechanical ventilation; increase in number or extension of areas of pulmonary consolidation. Assessment of radiological pattern and definition of the extent of lung involvement were performed and scored as previously described (Li et al., 2020). Patients with severe heart failure, bacterial infection, hematological neoplasm, severe neutropenia or thrombocytopenia, and serum alanine aminotransferase > 5x UNL were excluded. All enrolled subjects provided written informed consent. The study was approved by the Regional Institutional Review Board (Comitato Etico Regione Marche) and was conducted in accordance with the Declaration of Helsinki. A Simon’s optimal two-stage design was adopted. TCZ was administered as a single-dose intravenous infusion of 8 mg/kg over a 90-minute time span after premedication with 40 mg methylprednisolone. Patients were classified as responders (R) if fulfilling one of the following criteria: i) improvement of oxygen saturation by more than 3% points and/or increase in P/F by 50% and/or P/F increase above 150 mmHg 72 hours after tocilizumab AND persistence of this improvement at day 7, OR ii) no evidence of worsening of respiratory function as defined above. Dead and intubated patients were classified as Non-responders (NR). Samples of serum were collected at baseline (T0) and after 72 hours (T1) from TCZ infusion. Twenty-nine age- and gender-matched healthy control subjects (CTR), without SARS-CoV-2 infection or any other acute or chronic illness, were enrolled. Mean age of CTR was 64.1±8.6 years. Serum MiRNAs were quantified by RT-PCR and Droplet Digital PCR (ddPCR) in two independent laboratories as previously described (Ferracin et al., 2015; Mensà et al., 2019). To achieve normal distribution, results were expressed as log2(relative expression) and log2(copies/μl), respectively. Results were presented as Z-scores to allow proper comparison between relative and absolute quantification.

Data were tested for normality using the Shapiro-Wilk test. Student’s *t* test for independent samples and two-way mixed ANOVA with Tukey post hoc tests were used to compare the levels of investigated miRNAs between COVID-19 patients and CTR and between time points in COVID-19 patients, respectively. The agreement between RT-PCR and ddPCR was assessed using Pearson’s correlation and Bland-Altman analysis. Pearson’s coefficient was used for the estimation of the correlations between miRNAs expression levels and clinical parameters. Exploratory factor analysis was performed as previously described (Spazzafumo et al., 2013) to identify underlying factors in the study population. Two-tailed p value < 0.05 was considered significant. Statistical analysis was performed using SPSS version 26 (IBM, USA).

## 3. RESULTS

The final study case set was composed of 29 serum samples from patients enrolled in the clinical trial NCT04315480, as one serum sample was excluded due to hemolysis. Demographic and clinical characteristics of the samples are reported in **Table 1**. None of the patients were smokers. The median time between onset of symptoms and TCZ infusion was 9 days (IQR 4-14 days). At the end of the study, 16 patients were classified as responders (R) and 13 patients as non-responders (NR). Given the age-dependent expression of the investigated miRNAs (Mensà et al., 2019; Rusanova et al., 2018), analyses were conducted after controlling for age. A significant interaction between time and responder status was found for miR-146a-5p levels (F[1,26]=6.904, p=0.014, partial η^2^=0.210).

**TABLE 1.** Baseline clinical and demographic characteristics of 29 COVID-19 patients treated with tocilizumab (TCZ), divided into responders (R) and non-responders (NR). Data are mean (SD). P value from *t* tests for continuous variables and *z* tests for categorical variables. LMWH, low-molecular weight heparin.

| Variable | Responder (n=16) | Non-responder (n=13) | P value |
| --- | --- | --- | --- |
| Age (years) | 65.9 (10.6) | 69.4 (12.8) | 0.424 |
| Gender (males, %) | 11 (68.8%) | 6 (46.2%) | 0.219 |
| Time between onset of symptoms and TCZ infusion (days) | 9.8 (5.9) | 9.6 (3.0) | 0.925 |
| Concomitant treatments |  |  |  |
| Lopinavir-ritonavir or darunavir-cobicistat | 13 (81.3%) | 10 (76.9%) | 0.775 |
| Antibiotics | 10 (62.5%) | 9 (69.2%) | 0.705 |
| Prophylactic LMWH | 10 (62.5%) | 6 (46.2%) | 0.379 |
| IL-6 (pg/mL) | 33.1 (40.9) | 146.1 (252.4) | 0.088 |
| Hemoglobin (g/dL) | 12.9 (1.7) | 12.6 (1.1) | 0.604 |
| Neutrophils (n/mm<sup>3</sup>) | 5031 (2580) | 6382 (3708) | 0.258 |
| Lymphocytes (n/mm<sup>3</sup>) | 673 (262) | 650 (227) | 0.801 |
| Platelets (n/mm<sup>3</sup>) | 200063 (83781) | 217769 (76300) | 0.561 |
| D-dimer (ng/mL) | 573.1 (687.4) | 1109.4 (1498.4) | 0.287 |
| C-reactive protein (ng/mL) | 8.3 (5.3) | 13.2 (8.5) | 0.095 |
| PaO<sub>2</sub>/FiO<sub>2</sub> | 146.1 (75.4) | 162.2 (48.5) | 0.570 |
| 1-week follow-up (n, %) |  |  |  |
| Home discharge | 16 (100%) | 7 (53.8%) | 0.002 |
| Intensive care | 0 | 1 (7.7%) | 0.258 |
| Death | 0 | 5 (38.5%) | 0.006 |

Analysis of simple main effects of time revealed a significant increase in miR-146a-5p levels in R patients 3 days after the administration of TCZ (Z-score difference = 1.25; p<0.001), while no significant change was shown in NR patients (p=0.125). No significant differences in baseline miR-146a-5p levels were found between R and NR (p=0.392), while post-treatment miR-146a-5p levels were higher in R vs. NR (Z-score difference = 0.98; p=0.007) (**Figure 1A**). Notably, droplet digital ddPCR analysis, which allows for the quantification of miRNA copies/μl of serum, confirmed RT-PCR results (F[1,25]=5.696, p=0.025, partial η^2^=0.186), with a strong Pearson’s correlation between techniques (Pearson’s r=0.74, p<0.0001). The Bland-Altman agreement analysis revealed a small +0.32 Z-score unit bias [95% CI: −1.09 – 1.73] between ddPCR and RT-PCR, confirming a reasonable agreement between the two methods (**Figure 1B**). Absolute quantification of miR-146a-5p copies revealed a mean increase from 3.2±1.4 to 5.3±1.3 copies per μl in R patients and a mean decrease from 3.4±1.7 to 2.1±1.6 copies per μl in NR patients.

**Figure 1.**
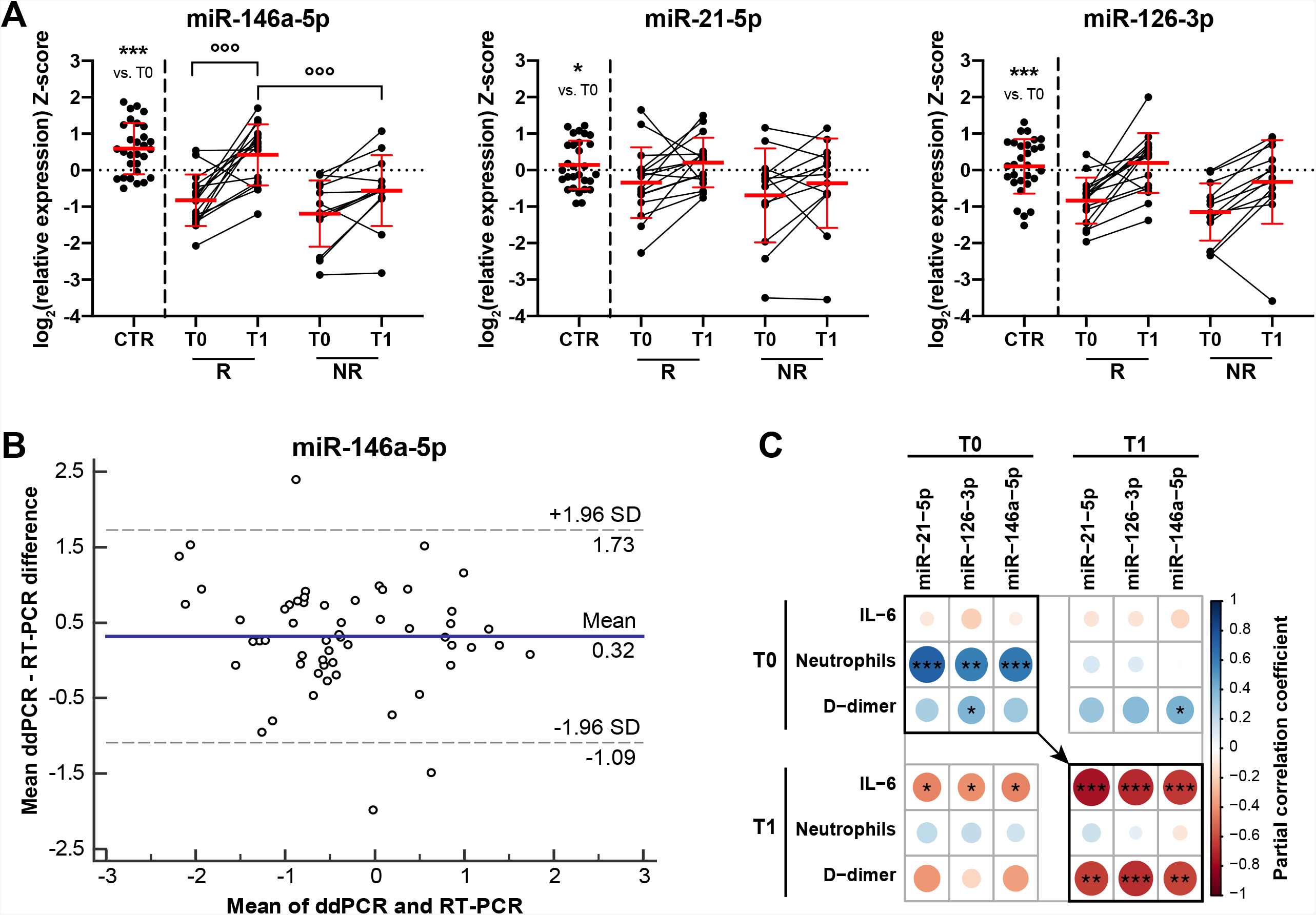
**(A)** Serum levels of miR-146a-5p, -21-5p, and -126-3p in 29 COVID-19 patients at baseline (T0) and after 72 hours from treatment with tocilizumab (T1), divided into responders (R) and non-responders (NR), and in 29 age-matched healthy control subjects (CTR). Data are expressed as Z-scores of log2(relative expression) and presented as mean ± SD. *, p<0.05; ***, p<0.001 for unpaired *t* test (CTR vs. COVID-19). °°°, p<0.001 for simple main effects of time (T0 vs. T1) and responder status (R vs. NR). **(B)** Bland-Altman plot for inter-method agreement between Droplet Digital PCR (ddPCR) and RT-PCR in the quantification of circulating miR-146a-5p. The blue line represents the mean bias between the two methods, the dashed lines indicate the limits of agreement. **(C)** Correlation plot showing partial correlations, controlling for age, between inflamma-miR levels and selected variables at both time points. Bold squares indicate correlations between variables assessed at the same time point. The color and the size of the circles depend on the magnitude of the correlation. Blue, positive correlation; red, negative correlation. Significant correlations are marked with * (p<0.05), ** (p<0.01), or *** (p<0.001).

No significant interaction between the responder status and time was showed for miR-21-5p (F[1,26]=1.089, p=0.306, partial η^2^=0.040). However, the main effect of time showed that circulating miR-21-5p was 0.433 SD higher at T1 compared to T0 (F[1,26]=5.048, p=0.033, partial η^2^=0.163), regardless of the responder status (**Figure 1A**). Notably, the main effect of time was confirmed by ddPCR (p=0.007). Bland-Altman analysis revealed a strong agreement between methods (ddPCR – RT-PCR Bias= −0.12, 95% CI: −0.26 – 0.03). TCZ treatment does not modulate angio-miR-126-3p levels (F[1,26]=0.621, p=0.438, partial η^2^=0.023), and no difference was observed between R and NR (main effect of responder status, p=0.202), or time points (main effect of time, p=0.266). No significant gender-related difference in the pre- and post-treatment levels of miR-146a-5p, miR-21-5p, and miR-126-3p were shown (data not shown).

Multiple comparisons of age-adjusted circulating inflamma-miRs showed significantly lower miR-146a-5p (mean difference = −1.498, p<0.001), miR-21-5p (mean difference = −0.486, p=0.025), and miR-126-3p (mean difference = −0.972, p<0.001) levels in COVID-19 patients compared to CTR subjects (**Figure 1A**).

Partial bivariate correlations adjusted for age were computed to analyze the relations between inflamma-miRs and selected variables, including plasma IL-6, hemoglobin, neutrophil, lymphocyte and platelet count, D-dimer, and PaO2/FiO2 ratio in COVID-19 patients at both time points. The results, summarized in the correlation plot in **Figure 1C**, show significant positive correlations between the baseline neutrophil count and the three inflamma-miRs. Moreover, negative correlations exist between post-intervention IL-6 levels and both T0 and T1 inflamma-miR levels. The circulating levels of the three miRNAs are negatively related to D-dimer level at T1.

Finally, an exploratory factor analysis (EFA) was conducted on the COVID-19 study population to identify latent variables that could be associated with clinical outcomes. The analysis returned 5 factors that cumulatively explain 84.7% of the total variance. The first factor, which included the 3 inflamma-miRs and age, explained most of the total variance (31.3%) and was the only factor that was significantly associated with mortality at the univariate logistic regression (p=0.043, OR 0.07 [95% CI 0.01 – 0.91]). These data confirmed the notion that a latent variable encompassing low levels of circulating inflamma-miRs and increasing age is associated with mortality, to a higher extent than plasma IL-6. Results of the EFA are summarized in **Figure 2A** and reported in **Supplementary Table 1**. Notably, age-adjusted baseline levels of miR-21-5p (p=0.016), -126-3p (p=0.006), and -146a-5p (p=0.008) were significantly lower in 5 NR COVID-19 patients (2 males and 3 females) who died in the 1-week follow-up period compared to NR survivors **(Figure 2B)**.

**Figure 2.**
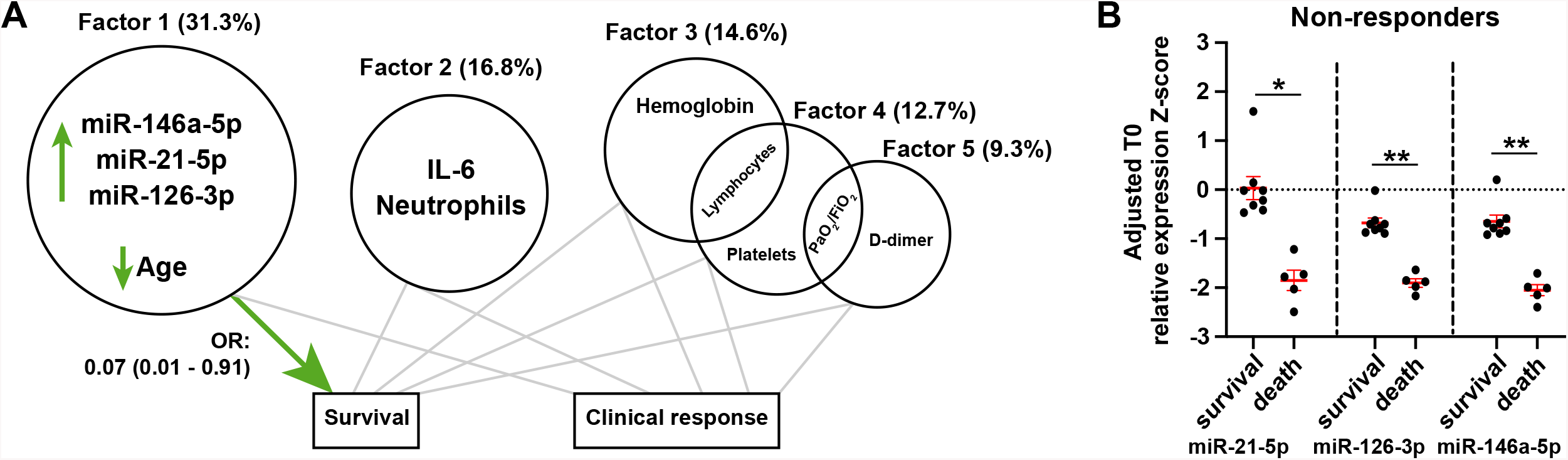
**(A)** Summary of exploratory factor analysis and subsequent logistic regressions on COVID-19 patients. Baseline variables are reported into each circle according to the factor loading. The areas of the circles are proportional to the amount of variance (reported in brackets) explained by each factor. Overlapping circles include variables loading onto two factors. The green arrow points out the significant association between factor 1 and survival, while the gray lines indicate non-significant associations. **(B)** Age-adjusted baseline miR-21-5p, -126-3p, and -146-5p levels in dead vs. survivor NR patients. Data are expressed as Z-scores of log2(relative expression) and presented as estimated marginal mean ± SEM. *, p<0.05; **, p<0.01 for one-way ANCOVA.

## 4. DISCUSSION

To our knowledge, this is the first report showing a significant association between serum miR-146a-5p levels and response to TCZ treatment in COVID-19 patients. Two different PCR-based assays, independently performed in two separate laboratories and providing relative and absolute quantifications of serum miR-146a-5p, revealed a significant increase of miR-146a-5p serum levels only in patients classified as “responders” to TCZ treatment. Although miR-146a-5p does not directly target the mRNAs of IL-6 or its receptor, miR-146a^−/−^ mice displayed high serum IL-6 levels, supporting the functional link between miR-146a-5p and IL-6 levels (Boldin et al., 2011). Indeed, miR-146a-5p acts as a negative regulator of NF-κB, which is in turn a widely acknowledged transcription factor of the IL-6 gene (Su et al., 2020). Also, the transcription of miR-146a-5p is under the control of NF-κB, even if the two molecules – IL-6 and miR-146a-5p – play opposite roles in the inflammatory process. A number of studies was focused on the complex regulatory mechanisms of IL-6 transcription, identifying not only NF-κB but also AP-1, sp-1 and NF-IL6-C/EBP as involved in such modulation (Akira et al., 1990; Libermann and Baltimore, 1990; Luo and Zheng, 2016). Therefore, the unbalance of the IL-6/miR-146a-5p axis could depend, at least in part, from IL-6-stimulating nuclear factors other than NF-κB or synergistically acting with NF-κB, thus dramatically exacerbating IL-6 synthesis without a concomitant induction of miR-146a-5p transcription. COVID-19 patients showed increased IL-6 levels and reduced miR-146a-5p levels compared to healthy age-matched subjects, pointing at the imbalance of the IL-6/miR-146a-5p physiological axis in the pathogenesis of SARS-CoV-2 infection. Noteworthy, a similar scenario was reported in the context of sepsis (Benz et al., 2016).

Overall, our data showed that the response to TCZ is associated with a significant reduction of plasma IL-6 and a significant increase of miR-146a levels, thus suggesting that TCZ treatment is able, at least in the set of responder patients, to offset the IL-6/miR-146a-5p axis unbalance. The observation that miR-146a-5p is significantly reduced in the group of non-responder COVID-19 patients with the worst outcome strongly supports this hypothesis. Notably, the inter-individual variability in the response to an anti-inflammatory treatment, i.e. TCZ, can be related to a number of different factors, including the genetic make-up, lifestyle, previous stimulations of immune cells, and the burden of senescent cells – mainly immune and endothelial cells.

Therefore, even though the number of patients is relatively low, our results suggest that low levels of circulating miR-146a-5p in COVID-19 patients may predict poor outcome among those who develop systemic hyper-inflammation. Notably, circulating miR-146a levels significantly decline with age and chronic age-related diseases and conditions, such as type 2 diabetes and frailty (Mensà et al., 2019; Ong et al., 2019). Current literature agrees about its pivotal role in inflammaging and age-related diseases (Grants et al., 2020). We recently suggested that the features of the current pandemic strongly suggest a role of inflammaging in worse COVID-19 outcomes, which mainly affect old people affected by age-related diseases (Bonafè et al., 2020; Storci et al., 2020). Interestingly, a randomized controlled trial evaluating the efficacy of a triple antiviral therapy with combined interferon beta-1b, ribavirin, and lopinavir-ritonavir on COVID-19 patients showed that the interferon-induced decline in circulating IL-6 is associated with earlier viral clearance (Hung et al., 2020). Also, glucocorticoids improve severe or critical COVID-19 by activating angiotensin-converting enzyme 2 (ACE2) and by reducing IL-6 levels (Xiang et al., 2020). It has been recently demonstrated that a strategy including high-dose methylprednisolone, followed by TCZ treatment if needed, may accelerate respiratory recovery, lower hospital mortality and reduce the likelihood of invasive mechanical ventilation in COVID-19-associated cytokine storm syndrome (Ramiro et al., 2020).

The data herein reported support the hypothesis that COVID-19 clinical progression may be accelerated by inflammaging, and/or that COVID-19 may accelerate inflammaging itself. Therefore, our results represent a proof-of-principle for the feasibility of dosing selected inflamma-miRs, specifically miR-146a-5p, as biomarkers of response to anti-inflammatory therapeutic intervention in COVID-19 patients. Moreover, such markers could represent themselves putative druggable targets (Su et al., 2020).

Regarding miR-21-5p and miR-126-3p, even if they were significantly reduced in COVID-19 patients compared to healthy subjects, no significant modulation was observed after TCZ treatment. Functional studies of miR-21 during inflammation are complicated by its numerous molecular targets and by the fact that its expression is regulated by multiple transcription factors (Sheedy, 2015). The existing evidences illustrate the role of miR-126-3p in inflammation, by regulating the NF-κB inhibitor Iκ-Bα (Feng et al., 2012), and endothelial activation (Zhang et al., 2020). However, a clear mechanism linking miR-126-3p and IL-6 levels was not yet described.

Notably, in the factor analysis only a latent variable encompassing all the three selected miRNAs and age showed to be predictive for survival after TCZ treatment.

In conclusion, further larger, multi-center, and possibly prospective clinical trials are necessary to identify the true potential of inflammamiR assessment as a predictive or prognostic biomarker in tocilizumab administration to COVID-19 patients and therefore validate the clinical relevance of our preliminary observations.

## Data Availability

The data that support the findings of this study are available from the corresponding authors upon reasonable request.

## Acknowledgments

The authors want to thank Azienda Ospedaliera Universitaria “Ospedali Riuniti”, Ancona, Italy for the support.

## Author contribution

JS performed statistical analysis, drafted the paper, and prepared the figure. Angelica Giuliani, GM, SL performed RNA extraction and RT-PCR measurements. NL and MF performed ddPCR quantification. GP and AF provided patient data and clinical advice. SSB, MP, and MM provided sera samples and laboratory data of COVID-19 patients. Armando Gabrielli is the principal investigator of the original trial. ADP critically revised the paper. MF, MB, and FO conceived the idea and wrote the paper. All authors critically revised the paper for important intellectual content and approved of the final version.

