## Supplementary Table 1 for "Decreased serum levels of the inflammaging marker miR-146a are associated with clinical response to tocilizumab in COVID-19 patients"

**Supplementary Table 1.** Results of the exploratory factor analysis (EFA) on the available variables evaluated at baseline. Varimax rotation converged in 6 iterations and yielded 5 factors with an eigenvalue > 1. Component loading >0.30 or <-0.30 are reported. Multiple univariate logistic regressions were computed to predict the association between each factor and the likelihood of adverse clinical response or death in TCZ-treated COVID-19 patients. B, beta value; OR, odds ratio; SE, standard error.

| | **Factor loading matrix** | | | | | |
|---|---|---|---|---|---|---|
| **Variables at T0** | **1** | **2** | **3** | **4** | **5** | **Cumulative** |
| **miR-146a-5p** | 0.975 | | | | | |
| **miR-21-5p** | 0.972 | | | | | |
| **miR-126-3p** | 0.946 | | | | | |
| **Age** | -0.738 | | | | | |
| **IL-6** | | 0.853 | | | | |
| **Neutrophils** | | 0.843 | | | | |
| **Hemoglobin** | | | 0.858 | | | |
| **Lymphocytes** | | | 0.829 | 0.34 | | |
| **Platelets** | | | | 0.915 | | |
| **D-dimer** | | | | | 0.918 | |
| **PaO2/FiO2** | | | | 0.539 | 0.664 | |
| **Eigenvalue** | 3.45 | 1.85 | 1.61 | 1.39 | 1.02 | |
| **Variance explained (%)** | 31.3 | 16.8 | 14.6 | 12.7 | 9.3 | 84.7 |
| **Logistic regression** | | | | | | |
| **Non-responder vs Factor** | | | | | | |
| B (SE) | -0.32 (0.44) | 0.59 (0.75) | -0.01 (0.47) | 0.11 (0.44) | 0.40 (0.42) | |
| OR (95% CI) | 0.74 (0.32 - 1.76) | 1.79 (0.41 - 7.78) | 0.99 (0.40 - 2.47) | 1.12 (0.47 - 2.65) | 1.49 (0.65 - 3.41) | |
| p | 0.501 | 0.435 | 0.990 | 0.799 | 0.349 | |
| **Death vs Factor** | | | | | | |
| B (SE) | -2.63 (1.30) | 0.48 (1.00) | -0.55 (0.78) | -0.22 (0.63) | -1.53 (2.07) | |
| OR (95% CI) | 0.07 (0.01 - 0.91) | 1.61 (0.23 - 11.34) | 0.58 (0.13 - 2.66) | 0.98 (0.29 - 3.35) | 0.22 (0.00 - 12.60) | |
| p | **0.043** | 0.633 | 0.483 | 0.972 | 0.461 | |
